# Psychometric Evaluation of the Japanese Yale-Brown Obsessive Compulsive Scale Modified for Olfactory Reference Disorder (ORD-YBOCS-J)

**DOI:** 10.1101/2024.08.19.24312169

**Authors:** Nanako Sano, Jennifer L. Greenberg, Saran Yoshida

## Abstract

**Objective:** The Yale-Brown Obsessive Compulsive Scale Modified for Olfactory Reference Syndrome (ORS-YBOCS) is a self-rated measure for assessing the severity of current olfactory reference disorder (ORD) symptoms. This scale has been translated into multiple languages and used in previous studies. However, its psychometric properties have not been investigated. The present study aimed to develop a Japanese ORS-YBOCS (newly named the ORD-YBOCS) and examine its reliability and validity.

**Methods:** A total of 104 individuals with self-reported ORD symptoms were included (mean age = 30.20 years, *SD* = 10.25). Internal consistency and factor structure were assessed by calculating Cronbach’s alpha and conducting confirmatory factor analysis (CFA) respectively. Convergent and divergent validity were examined via correlations with measures of Jikoshu-Kyofu symptoms, body image concerns, depression, and fear of negative evaluation.

**Results:** The Japanese ORD-YBOCS had good internal consistency (*α* = .81) and acceptable construct validity (*r* = .43 with Jikoshu-Kyofu symptoms, *r* = .20 with body image concerns, *r* = .56 with depression, and *r* = .27 with fear of negative evaluation). The scree plot suggested a one-factor model, but the CFA results did not support this (CFI = 0.76, TLI = 0.70, RMSEA = 0.14, and SRMR = 0.10).

**Conclusion:** This is the first study to examine the psychometric properties of the self-report ORS-YBOCS. The linguistically validated Japanese ORD-YBOCS showed good internal consistency and acceptable validity. These results suggest that the Japanese ORD-YBOCS is a valid and reliable tool to measure ORD symptom severity.

## 1. Introduction

Olfactory reference disorder (ORD) is an understudied obsessive-compulsive and related disorder characterized by a persistent preoccupation with emitting offensive odor, although the odor is unnoticeable or only slightly noticeable to others [1]. Individuals with ORD often engage in repetitive and excessive behaviors, such as asking for reassurance and excessive showering, to try to prevent, check, or hide the perceived odor. In previous literature, ORD was known as olfactory reference syndrome (ORS) or the Japanese “Jikoshu-Kyofu,” and its symptoms were classified respectively as either an example of a delusional disorder, somatic type or as a culture-bound syndrome under Taijin-Kyofusho in the Diagnostic and Statistical Manual of Mental Disorders (DSM-Ⅳ-TR) [2]. DSM-5 first mentioned ORS and Jikoshu-Kyofu together as “other specified obsessive-compulsive and related disorder” in its newly classified obsessive-compulsive and related disorders category [3]. The term ORD was established with the most recent publications of the International Classification of Diseases, Eleventh Revision (ICD-11) [4] and the DSM -5 TR [1].

Previous studies suggest that approximately 2% of the population is affected by ORD [5–7] and its symptoms can have a significant negative impact on an individual’s mental health as well as social, work, and daily activities [8,9]. Given the substantial psychosocial impact and the chronic course associated with ORD [8,9], there is a critical need to identify and treat individuals with this disorder. Yet, research in ORD remains scarce, with no randomized controlled trials on ORD published to date.

Research on ORD has been stymied in part by the lack of standardized assessment. Validated measures of ORD symptoms are needed to accurately assess its prevalence and response to treatment. Greenberg et al. developed a self-rated version of the Yale-Brown Obsessive Compulsive Scale Modified for Olfactory Reference Syndrome (ORS-YBOCS) [8]. This scale assesses the past week severity of ORD symptoms and was adapted from the self-rated version of the Yale-Brown Obsessive Compulsive Scale (Y-BOCS) [10–12] and the semi-structured interview version of the Yale-Brown Obsessive Compulsive Scale Modified for Body Dysmorphic Disorder (BDD-YBOCS) [13]. The ORS-YBOCS is comprised of 12 items assessing obsessions (item 1–5), compulsions (item 6–10), insight (item 11), and avoidance (item 12). Each item is rated on a Likert scale from 0–4, with a total score range of 0-48. Higher scores indicate greater severity. A total score of ≥ 20 on the ORS-YBOCS is used as a clinical cut-point to determine the presence of ORD, consistent with the widely used cut-point of ≥ 20 on the BDD-YBOCS that corresponds to a diagnosis of BDD [8,14].

The ORS-YBOCS self-rated version is easy to administer and enhances the feasibility of conducting studies with large sample sizes [7,15] and repeated measurements [16]. Moreover, self-report can reduce social desirability bias, particularly around sensitive information, and may help individuals gain a deeper understanding of their symptoms [17]. However, the psychometrics of the ORS-YBOCS have not been established and there is currently no Japanese measure of ORD symptoms.

The present study aimed to develop a Japanese ORS-YBOCS self-rated version and to examine its psychometric properties. First, we developed the Japanese ORS-YBOCS with its name changed to Japanese ORD-YBOCS (ORD-YBOCS-J) since ORD has been described in the ICD-11 [4] and DSM-5 TR [1] as a unique clinical term. These modifications were made with the permission of the original author of the ORS-YBOCS (i.e., the second author of the present study). Second, we examined the internal consistency of the instrument. We then examined the factor structure by performing confirmatory factor analysis. Lastly, we assessed the convergent and divergent validity of the ORD-YBOCS-J via correlations with measures of Jikoshu-Kyofu symptoms, body image concerns, depression, and fear of negative evaluation. With regard to convergent validity, we hypothesized a strong positive correlation between the ORD-YBOCS-J and Jikoshu-Kyofu items in the Egorrhea-experience Questionnaire (EQ) [18] as Jikoshu-Kyofu and ORD are thought to represent the same clinical entity [19]. For divergent validity, correlations between the ORD-YBOCS-J and measures of body image concern, depression, and fear of negative evaluation were expected to be weak or absent. Although researchers have recognized many similarities between ORD and BDD [20,21], and they can be comorbid (in one study, 30% of ORD patients had comorbid BDD [9]), there are important differences, including the focus of concern (odor in ORD, appearance in BDD) and poorer insight in those with ORD vs. BDD [20,22,23]. Similarly, depression and social anxiety (including fear of negative evaluation) are commonly comorbid to ORD but represent unique constructs that have been shown previously to have a weak correlation with ORD severity [7–9,24].

## 2. Methods

### 2.1 Participants

Eligibility criteria of the participants were adapted from Greenberg et al. [8,21]. Inclusion criteria were: (a) age of 18 years or older, (b) proficiency in Japanese, and (c) Answering “yes” to both of the following two questions: (1) Are you very worried about how you smell? (2) Do you spend a lot of time thinking about your odor concerns and wish you could think about them less? Exclusion criteria included: (a) diagnosis of disease or symptoms that can cause malodor and (b) diagnosis of schizophrenia. Of the 113 respondents, those diagnosed with a disorder that could potentially emit malodor (*n* = 7) or schizophrenia (*n* = 1) and those with detected inaccuracies in response to the Directed Questions Scale (*n* = 1) were excluded. Thus, 104 participants were included in the analysis (response rate: 92.04%). The sample size was designed to be at least 100 based on the requirements of the COSMIN Checklist [25].

### 2.2 Procedures

Online surveys were conducted using Google Forms between November and December 2023. Participants were recruited through social networking sites and by posting flyers in support groups for individuals with odor concerns. When answering the questions, the participants were instructed to think of their odor, including body odor or halitosis. The questions were designed such that only those who met the inclusion criteria and did not meet the exclusion criteria could proceed further. Participants were fully informed of the purpose of the study and their right to drop out of the survey at any time before submitting their responses. All participants were asked for consent to participate in the study, and only those who consented proceeded further. Individuals did not receive compensation for their participation. All procedures were approved by the Tohoku University Kawauchi-Minami Area “Medical and Health Research Involving Human Participants” Ethics Committee (2023-009).

### 2.3 Materials

#### 2.3.1 Demographic Information

Participants were asked to report age, gender, highest level of education, employment status and marital status. Participants were also asked: (1) body parts of concern [8] and (2) the frequency of experiences in which others directly indicated that the participant’s odor was offensive.

#### 2.3.2 Translation Procedures for the Japanese ORD-YBOCS

First, the original ORS-YBOCS [8] was translated into Japanese by a native Japanese speaker with fluency in English. We then compared a forward translation of the ORS-YBOCS with the Japanese Y-BOCS [17] and reconciled with them, given the similarity between the original self-report Y-BOCS [12] and ORS-YBOCS [8]. This reconciliation process was done with an involvement of the professional psychologist who has extensive experience working with patients with obsessive-compulsive disorder (OCD) in a clinical setting. Next, the forward translation was back-translated by a Japanese-English bilingual assistant professor in the Department of Clinical Psychology who was not involved in the forward-translation process. The back-translation was sent to the original author of the ORS-YBOCS to evaluate the equivalence of the scales. Finally, we conducted a cognitive debriefing of the ORD-YBOCS-J: five native Japanese speaking graduate students completed the ORD-YBOCS-J and provided some additional minor language modifications to improve understanding of the items.

The research team (JG, NS, SY) involved in the forward translation, consolidation, and backward translation processes then evaluated the final version of the ORD-YBOCS-J and confirmed the equivalence between the Japanese and English versions. The translation processes were based on the guidelines proposed by the ISPOR Task Force for the translation and cultural adaptation process of the patient-reported outcomes [26].

In developing the Japanese version, we intended to reflect the conceptual meaning of the original version rather than adhere to a literal translation. In addition, several modifications were made to make it easier for the Japanese to respond, including (1) adding instructions from the Japanese Y-BOCS [17] to items 1 and 6 that ask participants to select “0” for the subsequent four items if they chose 0 on item 1 or 6 with permission of the first translator of the Japanese Y-BOCS, (2) modifying the original term “body odor” to “odor” to assess all types of odor and to be consistent with the diagnostic criteria of the ICD-11 [4] that includes halitosis as one of the odor concerns, and (3) excluding two examples of “airing out the area” in the item 6 check list that might be irrelevant or difficult to understand for a Japanese population (i.e., “getting up” and “taking off shoes”).

As in the original version, the ORD-YBOCS-J contains 12 items, each scored on a scale of 0–4. Items 2 (interference due to obsessions) and 6 (time spent in repetitive and excessive behaviors) also include checklist items to better to understand the range of these specific symptoms. However, these checklist items are not included in the total score. The total score ranges from 0 to 48, with higher scores indicating greater severity of ORD symptoms.

#### 2.3.3 Qualitative Participant Feedback on the ORD-YBOCS-J

Two open-ended questions were asked to elicit participants’ feedback on the measure: one asked respondents to share any difficulties in answering the ORD-YBOCS-J, and the other asked for general opinions or impressions of the scale.

#### 2.3.4 Measures to Evaluate Construct Validity

##### Egorrhea-experience Questionnaire (EQ)

The EQ contains 32 items that evaluate episodes of egorrhea-related symptoms, including those observed under normal conditions [18]. The EQ covers four dimensions: “Sekimen-Kyofu” (erythrophobia), “Jikoshu-Kyofu” (fear of self-odor), “Shukei-Kyofu” (dysmorphophobia), and “Shikou-Denpa” (thought broadcasting). Seven items that reflect the construct of “Jikoshu-Kyofu” are each rated on a scale from 1 to 5; the total score ranges from 7 to 35. Previous studies have shown good internal consistency in adolescents’ samples (*α* = .89-.90 [6,18]).

##### Japanese Body Image Concern Inventory (J-BICI)

The J-BICI comprises 19 items, each rated on a scale of 1–5 for measuring body image concerns [27,28]. The total score ranges from 19 to 95. Higher scores indicate more severe body image concerns. The J-BICI has good internal consistency (*α* = .91), high retest reliability (*r* = .88), and good concurrent validity (*r* = .77) [28].

##### Japanese Patient Health Questionnaire-9 (J-PHQ-9)

The J-PHQ-9 contains nine items assessing depressive symptoms for the past two weeks, each rated on a scale from 0 to 3 [29,30]. The total score ranges from 0 to 27. Higher scores indicated more severe depressive symptoms. Good internal consistency has been reported (*α* = .93) [31].

##### Japanese Short Fear of Negative Evaluation Scale (SFNE-J)

The SFNE-J includes 12 items, each rated on a scale of 1–5; the total score ranges from 12 to 60 [32,33]. It evaluates one’s fear of negative evaluation from others. A higher score indicates a higher fear of negative evaluations. All reverse items were inverted before calculating the total scores. The SFNE-J has good internal consistency and adequate validity [33,34].

#### 2.3.5 Directed Questions Scale

The Directed Questions Scale (DQS) [35] was applied to detect participants with problematic responses such as not reading carefully. An item mentioning “Here, select the bottom option ‘always true’ was placed within the J-BICI items as a DQS, referring to Shimizu et al. [36] and Maniaci and Rogge [35]. DQS has been reported not to influence scale validity [37].

### 2.4 Statistical Analysis

First, descriptive statistics were calculated for participants’ demographics and odor concern-related items. Second, descriptive statistics and item analysis of the ORD-YBOCS-J were conducted. The reliability of the ORD-YBOCS-J were then analyzed by computing Cronbach’s alpha value. A scree plot of the ORD-YBOCS-J was generated to explore the factor structure, and confirmatory factor analysis (CFA) was performed. Lastly, convergent and divergent validities were examined by calculating Pearson’s product-moment correlation coefficients between the measurements.

All analyses were performed using R version 4.2.2. In CFA, the model was considered good if the Comparative Fit Index (CFI) is .95 or higher, the Tucker-Lewis Index (TLI) is .95 or higher, the root mean square error of approximation (RMSEA) is .06 or lower, and the standard root mean square residual (SRMR) is .08 or lower [38]. For the correlation, .10 ≤ | *r* | < .30 is considered weak, .30 ≤ | *r* | < .50 is moderate, and above .50 is strong [39].

## 3. Results

### 3.1 Demographics and Odor Concerns Characteristics

Table 1 summarizes the demographic characteristics of participants. Participants were on average 30.20 (*SD*=10.25) years old and predominately (81.73%) female. The most common body parts of concern were the armpits (22.12%), followed by the mouth (21.15%), general body (19.23%), and anus (12.50%). Participants endorsed having 1 to 13 (*M* = 3.74) body parts of concern (*n* =101, excluding three respondents who could not identify the body parts, e.g., “I do not know which body parts are smelly”). Participants reported the frequency with which others pointed out the participant’s odor as being offensive as: never (18.27%), rarely (21.15%), sometimes (28.85%), often (24.04%), and always (7.69%).

**Table 1.**
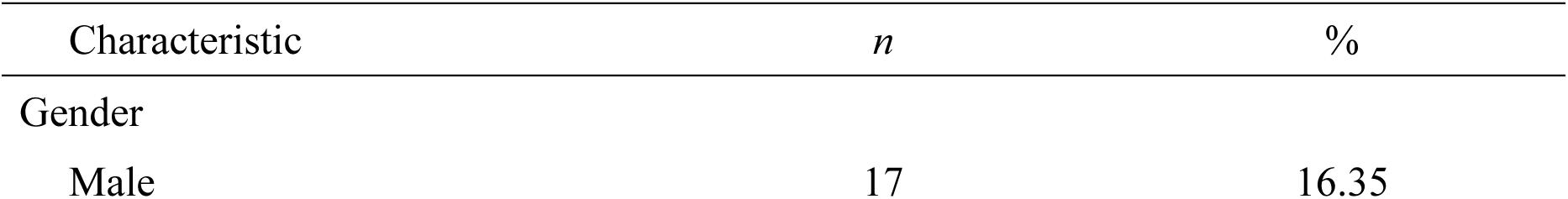
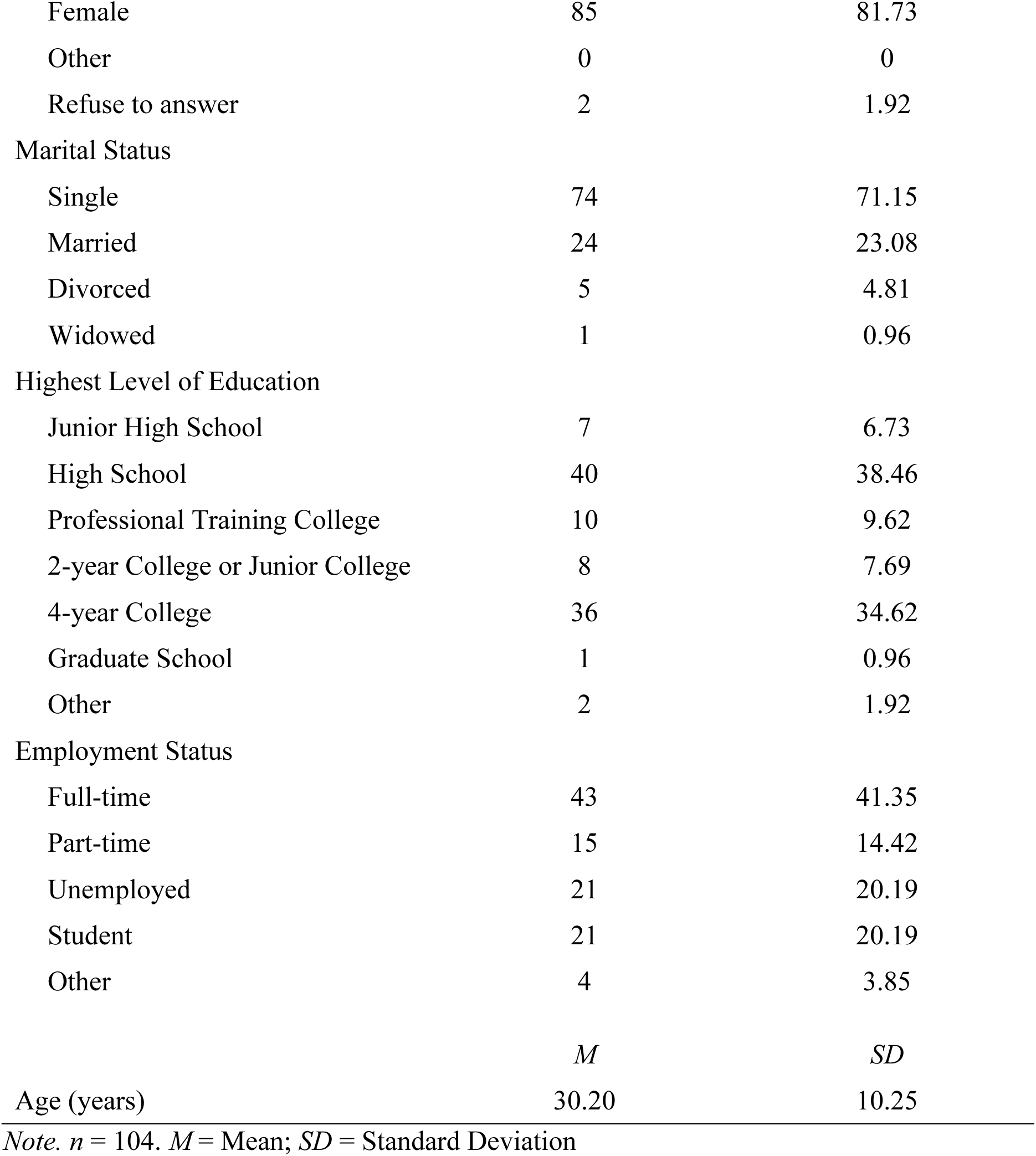
Demographic Characteristics of Participants.

*Demographic Characteristics of Participants*
| Characteristic | *n* | % |
| --- | --- | --- |
| Gender |  |  |
| Male | 17 | 16.35 |
| Female | 85 | 81.73 |
| Other | 0 | 0 |
| Refuse to answer | 2 | 1.92 |
| Marital Status |  |  |
| Single | 74 | 71.15 |
| Married | 24 | 23.08 |
| Divorced | 5 | 4.81 |
| Widowed | 1 | 0.96 |
| Highest Level of Education |  |  |
| Junior High School | 7 | 6.73 |
| High School | 40 | 38.46 |
| Professional Training College | 10 | 9.62 |
| 2-year College or Junior College | 8 | 7.69 |
| 4-year College | 36 | 34.62 |
| Graduate School | 1 | 0.96 |
| Other | 2 | 1.92 |
| Employment Status |  |  |
| Full-time | 43 | 41.35 |
| Part-time | 15 | 14.42 |
| Unemployed | 21 | 20.19 |
| Student | 21 | 20.19 |
| Other | 4 | 3.85 |
|  | *M* | *SD* |
| Age (years) | 30.20 | 10.25 |

Note. $n = 104$ . $M$ = Mean; $SD$ = Standard Deviation

### 3.2 Item Analysis and Reliability

The mean (*M*), standard deviation (*SD*), minimum (*Min*), maximum (*Max*), item-total correlation (*ITC /* I-T correlation), and Cronbach’s alpha value when the item was eliminated are shown in Table 2. Item means ranged from 2.07 to 3.27, indicating a range from “moderate” to “severe”. The I-T correlation was lower for items 4 (*r* = .12), 11 (*r* = .20), and 9 (*r* = .30). Correlations between items 4, 9, and 11 were as follows: *r* = .60 (item4–9), *r* = .04 (item 4–11), *r* = .14 (item 9–11). Cronbach’s alpha for all 12 items was .81, and the results suggest that removing items 4 and 11 improved Cronbach’s alpha by .02 and .01, respectively. The total score of the ORD-YBOCS-J ranged from 15 to 48, with 97.12% scoring 20 or higher. No floor or ceiling effects were observed. The total 12-item skewness was −0.22, and kurtosis was −0.35.

**Table 2.**
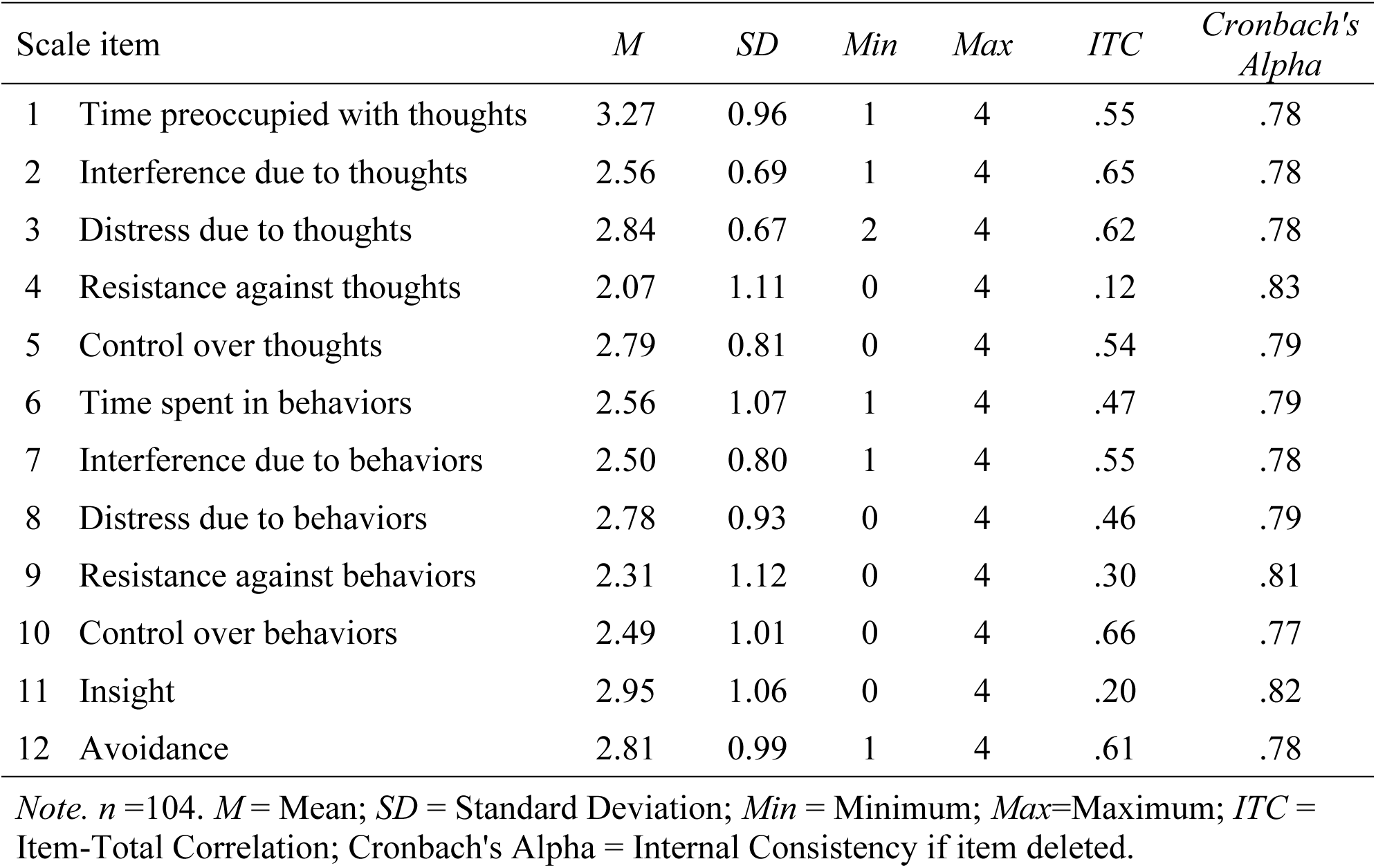
Item Statistics on the Japanese ORD-YBOCS.

| Scale item | *M* | *SD* | *Min* | *Max* | *ITC* | *Cronbach's Alpha* |
| --- | --- | --- | --- | --- | --- | --- |
| 1 Time preoccupied with thoughts | 3.27 | 0.96 | 1 | 4 | .55 | .78 |
| 2 Interference due to thoughts | 2.56 | 0.69 | 1 | 4 | .65 | .78 |
| 3 Distress due to thoughts | 2.84 | 0.67 | 2 | 4 | .62 | .78 |
| 4 Resistance against thoughts | 2.07 | 1.11 | 0 | 4 | .12 | .83 |
| 5 Control over thoughts | 2.79 | 0.81 | 0 | 4 | .54 | .79 |
| 6 Time spent in behaviors | 2.56 | 1.07 | 1 | 4 | .47 | .79 |
| 7 Interference due to behaviors | 2.50 | 0.80 | 1 | 4 | .55 | .78 |
| 8 Distress due to behaviors | 2.78 | 0.93 | 0 | 4 | .46 | .79 |
| 9 Resistance against behaviors | 2.31 | 1.12 | 0 | 4 | .30 | .81 |
| 10 Control over behaviors | 2.49 | 1.01 | 0 | 4 | .66 | .77 |
| 11 Insight | 2.95 | 1.06 | 0 | 4 | .20 | .82 |
| 12 Avoidance | 2.81 | 0.99 | 1 | 4 | .61 | .78 |

*Note.* $n = 104$ . *M* = Mean; *SD* = Standard Deviation; *Min* = Minimum; *Max* = Maximum; *ITC* = Item-Total Correlation; Cronbach's Alpha = Internal Consistency if item deleted.

### 3.3 Factor Analysis

A scree plot is presented in the Supplemental Appendix. The first eigenvalue accounted for over 30% of the total. Research in Item Response Theory suggests that when the first eigenvalue accounts for more than 20% of the total, it implies that there is unidimensionality [40]. Additionally, in the scree plot, the sharpest curve was observed from the first eigenvalue to the second. Therefore, it was proposed that a one-factor model may be appropriate for the ORD-YBOCS-J. The results of the CFA for the one-factor model are shown in Table 3 and suggest that items 4, 9, and 11 had low factor loadings. The fit for the one-factor model was also inadequate (CFI = 0.76, TLI = 0.70, RMSEA = 0.14, and SRMR = 0.10). However, we decided to adopt all 12 items from the original scale as the ORD-YBOCS-J for the following reasons: (a) *α* = .81, (b) neither floor nor ceiling effects were observed, (c) I-T correlation was weak on item 4, but other items were .20 or stronger, which was considered acceptable [41], (d) item 4 reflects an important aspect for evaluating ORD symptom severity and has clinical significance [11], and (e) the scree plot suggested the unidimensionality of the scale.

**Table 3.** Factor Loadings from the Confirmatory Factor Analysis.

| Scale item | *F* |
| --- | --- |
| 2 Interference due to thoughts | .79 |
| 7 Interference due to behaviors | .76 |
| 3 Distress due to thoughts | .76 |
| 12 Avoidance | .68 |
| 10 Control over behaviors | .68 |
| 5 Control over thoughts | .64 |
| 1 Time preoccupied with thoughts | .55 |
| 8 Distress due to behaviors | .55 |
| 6 Time spent in behaviors | .53 |
| 9 Resistance against behaviors | .20 |
| 11 Insight | .18 |
| 4 Resistance against thoughts | .01 |

*Note.* *F* = Factor Loadings

### 3.4 Convergent and Divergent Validity

The results of the descriptive statistics and correlation tests for the ORD-YBOCS-J, EQ, J-BICI, J-PHQ-9, and SFNE-J are presented in Table 4. For convergent validity, the correlation coefficient between the EQ and ORD-YBOCS-J was moderate (*r* = .43, *p* < .001). One-third of participants (33.65 %) scored the maximum score on the EQ (35 points), and their total scores of the ORD-YBOCS-J range from 27 to 48. For divergent validity, the correlation coefficients between the ORD-YBOCS-J and J-BICI (*r* = .20, *p* = .04) and between the ORD-YBOCS-J and SFNE-J (*r* = .27, *p* =.01) were weak, whereas the correlation coefficient between the ORD-YBOCS-J and J-PHQ-9 (*r* = .56, *p* < .001) was strong.

**Table 4.** Descriptive Statistics and Correlations for Construct Validity Measures.

| Measure | *M* | *SD* | *Range* | *Cronbach's Alpha* | *Correlations* |  |  |  |  |
| --- | --- | --- | --- | --- | --- | --- | --- | --- | --- |
|  |  |  |  |  | 1 | 2 | 3 | 4 | 5 |
| 1 ORD-YBOCS-J | 31.91 | 6.41 | 15-48 | .81 | — |  |  |  |  |
| 2 EQ (Jikoshu-Kyofu) | 32.36 | 3.45 | 15-35 | .81 | .43** | — |  |  |  |
| 3 J-BICI | 55.49 | 15.70 | 23-91 | .94 | .20* | .24* | — |  |  |
| 4 J-PHQ-9 | 14.97 | 7.03 | 1-27 | .88 | .56** | .30** | .36** | — |  |
| 5 SFNE-J | 48.38 | 9.75 | 12-60 | .92 | .27** | .33** | .47** | .34** | — |

*Note.* $n = 104$ . ORD-YBOCS-J = Japanese Yale-Brown Obsessive Compulsive Scale Modified for Olfactory Reference Disorder; EQ = Egorrhea-experience Questionnaire; J-BICI = Japanese Body Image Concern Inventory; J-PHQ-9 = Japanese Patient Health Questionnaire-9; SFNE-J = Japanese Short Fear of Negative Evaluation Scale. Correlations are Pearson product-moment correlation coefficient. *M* = Mean; *SD* = Standard Deviation.
\* $p < .05$ , \*\* $p < .01$ .

### 3.5 Participant Feedback on the ORD-YBOCS-J

One participant felt that in completing the ORD-YBOCS-J, “I understand that some individuals present with more severe symptoms than I do.” Another participant expressed disgust with being asked about insight (item 11). Thirteen participants reported some difficulty understanding scale items, including overall difficulty (*n* = 2), difficulty with items 4 and 9 (*n* = 4), item 11 (*n* = 2), item 6 (*n* = 1), and item 8 (*n* = 1), and unspecified (i.e., participants mentioning difficulty with a specific item, but did not identify which item they were referring to.) (*n* = 4) (the total *n* exceeded 13 because one participant reported difficulty with two items). In response to this feedback and to further enhance clarity, the authors subsequently made minor modifications to the text (i.e., adding “For example” between the first and the second sentences of item 4, with permission of the original author) and formatting (i.e., adding a line break before the parentheses in items 4, 6, 9, and 11).

## 4. Discussion

This study aimed to develop and linguistically validate a Japanese ORD-YBOCS and evaluate its psychometric properties. The item analysis results showed a weak I-T correlation only for item 4 (*r* = .12), which assessed one’s resistance against thoughts related to odor concerns. However, we decided to retain this item in the ORD-YBOCS-J. Goodman et al. [11], the developers of the Y-BOCS, noted that based on their clinical experience, patients with more severe OCD show less resistance. In addition, the results of the scree plot showed that the first eigenvalue accounted for over 30% of the total, and the calculation of the Cronbach’s alpha showed good internal consistency (*α* = .81), suggesting that it is reasonable to include all 12 items in the ORD-YBOCS-J.

The internal consistency of the ORD-YBOCS-J was good (*α* = .81). With regard to the convergent validity, the correlation between the EQ and ORD-YBOCS-J was moderate (*r* = .43), but not strong. This may imply that convergent validity in the self-reported ORD sample cannot be assessed using the EQ. Approximately 33.7% of participants scored at the maximum of the EQ total score range (35 points). Among those with an EQ score of 35, ORD-YBOCS-J total scores were in the moderate to extreme range (27-48), which may suggest that the ORD-YBOCS-J can assess severe symptoms with a good precision, but the EQ cannot, resulting in a weak correlation coefficient between two measurements. The EQ measures episodes of egorrhea-related symptoms and was developed and tested in adolescents, including egorrhea-related symptoms observed in a non-clinical sample [18]. Its reliability and validity have not been examined in adult or clinical samples. Thus, while the Jikoshu-Kyofu items in the EQ and ORD-YBOCS-J assess similar constructs, given its scale characteristics (i.e., developed and tested for adolescent, non-clinical samples), the EQ may not be an optimal measure of convergent validity for the ORD-YBOCS-J.

Divergent validity was supported to some extent. First, weak correlations were found between the ORD-YBOCS-J and J-BICI (*r* = .20) and between the ORD-YBOCS-J and SFNE-J (*r* = .27). However, the correlation between ORD-YBOCS-J and J-PHQ-9 was strong (*r* = .56). This finding is consistent with previous studies of the Y-BOCS or BDD-YBOCS, which have reported strong positive correlations between the Y-BOCS or BDD-YBOCS scores and depression scores [10,42]. A strong correlation between ORD symptom severity and depression has not been reported in the extant ORD literature. However, the ORD literature is scarce and it may be that the correlation between ORD symptom severity and depression becomes significant only at higher levels of ORD severity. Participants in the current study had more severe ORD symptoms overall than previously reported, for example in Greenberg et al. [8] (*t* (355) = 10.52, *p* < .001, 95%CI [4.80, 7.02]). Furthermore, Greenberg et al. [8] found that female participants reported more severe ORD symptoms than male participants, and the proportion of females in the current study (81.73%) was much higher than that in the Greenberg et al. [8] study (32.81%). Thus, differences in participant severity and gender may explain the differences in correlations between ORD symptom severity and depression that emerged in this study. Although divergent validity using depression was not successfully evaluated, our results suggest that the ORD-YBOCS-J has acceptable divergent validity.

The scree plot and good internal consistency suggested a one-factor model for the ORD-YBOCS-J. However, this was not supported by the results of the CFA (CFI = 0.76, TLI = 0.70, RMSEA = 0.14, and SRMR = 0.10). Items 4, 9, and 11 had low factor loadings (.01–.20) whereas the other items had moderate to high loadings (.53–.79). Items 4, 9, and 11 also showed lower I-T correlations (*rs* = .12–.30), while other items showed moderate to strong correlations (*rs* = .46–.66). These results can be interpreted in two ways: first, items 4, 9, and 11 might measure constructs other than ORD symptom severity, and second, these three items might be difficult for participants to respond to.

The possibility that items 4, 9, and 11 play a unique role relative to other items has been discussed in the areas of the Y-BOCS [43] and BDD-YBOCS [44]. In particular, OCD researchers have long discussed whether to remove resistance/control items, including item 4, from the Y-BOCS [43,45,46], and the Y-BOCS-Second Edition was developed with item 4 deleted [45]. Drawing on the work of previous studies [47,48], Storch et al. [45] has suggested that the stronger the resistance to obsessions, the more severe the impairment. In the present study, we chose to retain item 4 because of its potential clinical importance [11], but its inclusion warrants further investigation. The low loading of item 11 found in this study was also reported in a semi-structured BDD-YBOCS [44]. In addition, the insight item on the Y-BOCS was considered to measure a secondary aspect of OCD [43]. Given that the ICD-11 [4] has an insight specifier, item 11 on the ORD-YBOCS-J may also reflect a related, but distinct construct that would be more appropriately measured separately from the severity of ORD symptoms. Another possible explanation for the low factor loadings and I-T correlations for items 4, 9, and 11 is that these three items were difficult to answer for some participants. The role of items 4, 9, and 11 needs to be further examined with a reexamination of the factor structure of the ORD-YBOCS-J. Meanwhile, the factor structures of the Y-BOCS and BDD-YBOCS have also not yet reached consensus [13,16,42,44,49,50].

### 4.1 Participant Feedback on the Japanese ORD-YBOCS

Qualitative feedback from participants highlighted some of the benefits and risks of completing the self-report ORD-YBOCS-J. One participant described having a deeper understanding of their odor-related thoughts and behaviors after completing the ORD-YBOCS-J, which is consistent with a previous study which found that the self-administered Y-BOCS allowed patients to view their symptoms more objectively from their clinical experience in using the scale [17]. However, one participant in the present study reported that the insight item (item 11) was uncomfortable and even found it hard to believe it was included on the scale. Poor insight has been reported among individuals with ORD symptoms [9]. Directly pointing out unreasonable thoughts or behaviors may disgust them and make the treatment process more difficult [51]. Item 11 is included in the ORD-YBOCS-J because of the clinical importance of insight in ORD, with ICD-11 having an insight specifier [4]. However, as noted above, it is possible that insight may be better assessed with a separate, more nuanced, and/or clinician-administered measure. Further research with a larger sample size is needed to draw conclusions about the impact of responding to the ORD-YBOCS-J.

### 4.2 Limitations

The study has several limitations that must be considered. First, the sample consisted primarily of female participants. Therefore, our findings may not be generalizable to all with self-reported ORD symptoms. The high female to male ratio in the current study is in contrast to Greenberg et al. [8]. Previous studies conducted in Japan reported mixed results with regard to the gender ratio [52–54]. It is difficult to determine whether the predominantly female sample in this study reflects the true prevalence of ORD symptoms in Japan. Sex and gender differences should be further investigated in future studies.

The second limitation was the use of self-reporting to screen for ORD symptoms. Participants diagnosed with an illness that could potentially emit malodor were excluded from the study. Notwithstanding, those without a diagnosis but with a noticeable malodor might have been included since we did not assess participants’ odor objectively. Indeed, 31.73% of the participants selected either “often” or “always” when asked if/how often others had ever pointed out that the participant’s odor was offensive. It is not clear whether this may reflect a true malodor vs. ideas of reference commonly observed in ORD. We also excluded participants based on a self-reported diagnosis of schizophrenia; however, it is possible that the final sample included those with schizophrenia who had not received or were otherwise unaware of their diagnosis. Future studies should evaluate the psychometric properties of the ORD-YBOCS-J using an ORD sample screened by experienced clinicians or limited to those diagnosed with ORD. However, self-report assessment has the benefit of greater anonymity, making it easier for individuals with ORD symptoms, who often feel shame [55], to participate the survey, and can reduce social desirability bias. In addition, the use of self-report allowed individuals with ORD symptoms who are not connected to a clinic to participate in the present study. This is particularly beneficial given that ORD patients are known to rarely seek psychiatric care in Japan [51]. Self-report is also more efficient and cost-effective as it can often be completed quickly and independently without the need for a clinician, which enhances feasibility for engaging larger, more generalizable samples [8].

The third limitation is the sample size. The sample size in the present study was greater than 100, which is considered “very good” according to the COSMIN guidelines [25]. However, some researchers argue that 300 or more is appropriate for conducting factor analyses [56,57]. The poor model fit of the ORD-YBOCS-J in the CFA may be due to an insufficient sample size. The factor structure of the ORD-YBOCS-J needs to be re-evaluated with a sample size greater than 300. Lastly, based on participant feedback, some scale wording was modified slightly to enhance clarity. Thus, the final version of the ORD-YBOCS-J should be evaluated in future studies.

## 5. Conclusion

Standardized assessments of ORD are needed to better understand the prevalence and clinical impact of ORD as well as its response to treatment. This is the first study to examine the psychometric properties of the self-report ORS-YBOCS, which has been translated into multiple languages and used in previous studies. Although our findings should be interpreted cautiously, the results suggest that the linguistically validated Japanese ORD-YBOCS (ORD-YBOCS-J) has good internal consistency and acceptable validity. These results suggest that the Japanese ORD-YBOCS is valid and reliable tool to measure ORD symptom severity. More work is needed to replicate the psychometrics of the ORD-YBOCS-J, including its convergent and divergent validity, factor structure, sensitivity to treatment, re-test reliability, and validity of the cut-off score, using a clinical sample.

## CRediT authorship contribution statement

**Nanako Sano**: Writing-original draft, Conceptualization, Methodology, Data curation, Investigation, Formal analysis. **Jennifer L. Greenberg**: Writing-review & editing, Conceptualization, Resources, Validation. **Saran Yoshida**: Writing-review & editing, Conceptualization, Methodology, Supervision.

## Acknowledgments

The authors would like to express our gratitude to Dr. Lenna Schlemper for her support in the scale translation, and to MEXT/JSPS WISE Program : Advanced Graduate Program for Future Medicine and Health Care, Tohoku University for their support in this work.

## Data availability

Data will be made available on request. The language of the data will be Japanese only.

## Declaration of Generative AI and AI-assisted technologies in the writing process

During the preparation of this work, the first author used DeepL to improve readability and languages of the work. After using this tool, the authors reviewed and edited the content as needed and take full responsibility for the content of the publication.

## Supplemental Appendix

*Scree Plot for the Japanese ORD-YBOCS*

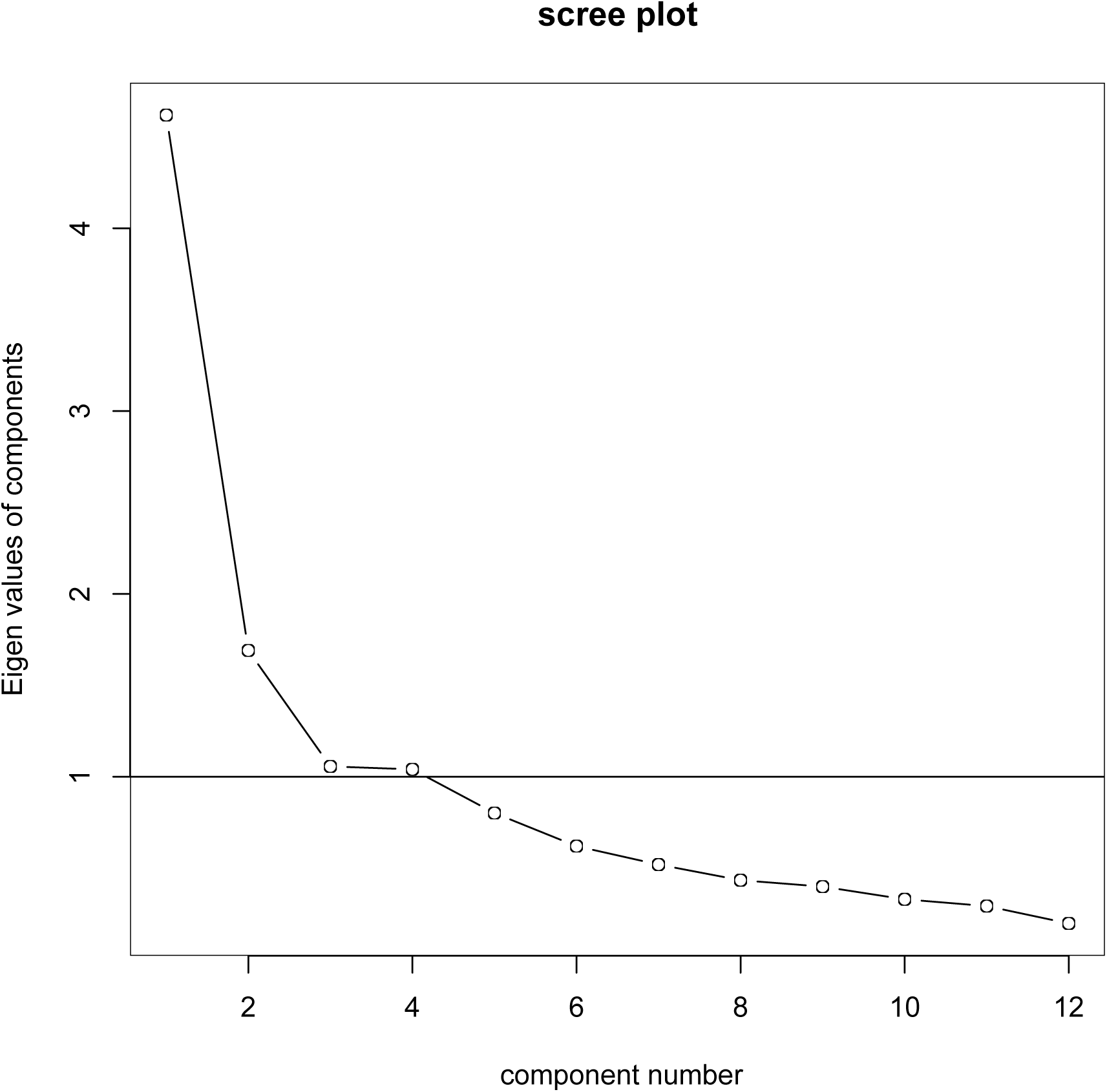

